# First-in-Human Trial of a SARS-CoV-2 Recombinant Spike Protein Nanoparticle Vaccine

**DOI:** 10.1101/2020.08.05.20168435

**Authors:** Cheryl Keech, Gregory M. Glenn, Gary Albert, Iksung Cho, Andreana Robertson, Patricia Reed, Susan Neal, Joyce S. Plested, Mingzhu Zhu, Shane Cloney-Clark, Haixia Zhou, Gale Smith, Nita Patel, Matthew B. Frieman, Robert E. Haupt, James Logue, Marisa McGrath, Stuart Weston, Pedro A. Piedra, Chinar Desai, Kathleen Callahan, Maggie Lewis, Patricia Price-Abbott, Neil Formica, Vivek Shinde, Louis Fries, Jason D. Lickliter, Paul Griffin, Bethanie Wilkinson

## Abstract

**Background:** NVX-CoV2373 is a recombinant nanoparticle vaccine composed of trimeric full-length SARS-CoV-2 spike glycoproteins. We present the Day 35 primary analysis of our trial of NVX-CoV2373 with or without the saponin-based Matrix-M1 adjuvant in healthy adults.

**Methods:** This is a randomized, observer-blinded, placebo-controlled, phase 1 trial in 131 healthy adults. Trial vaccination comprised two intramuscular injections, 21 days apart. Primary outcomes were reactogenicity, safety labs, and immunoglobulin G (IgG) anti-spike protein response. Secondary outcomes included adverse events, wild-type virus neutralizing antibody, and T-cell responses.

**Results:** Participants received NVX-CoV2373 with or without Matrix-M1 (n=106) or placebo (n=25). There were no serious adverse events. Reactogenicity was mainly mild in severity and of short duration (mean ≤2 days), with second vaccinations inducing greater local and systemic reactogenicity. The adjuvant significantly enhanced immune responses and was antigen dose-sparing, and the two-dose 5μg NVX-CoV2373/Matrix-M1 vaccine induced mean anti-spike IgG and neutralizing antibody responses that exceeded the mean responses in convalescent sera from COVID-19 patients with clinically significant illnesses. The vaccine also induced antigen-specific T cells with a largely T helper 1 (Th1) phenotype.

**Conclusions:** NVX-CoV2373/Matrix-M1 was well tolerated and elicited robust immune responses (IgG and neutralization) four-fold higher than the mean observed in COVID-19 convalescent serum from participants with clinical symptoms requiring medical care and induced CD4+ T-cell responses biased toward a Th1 phenotype. These findings suggest that the vaccine may confer protection and support transition to efficacy evaluations to test this hypothesis. (Funded by the Coalition for Epidemic Preparedness Innovations; ClinicalTrials.gov number, NCT04368988).

## Introduction

Coronavirus disease 2019 (COVID-19) has spread globally at a rapid pace since its inception in December 2019 in Wuhan, China and was declared a pandemic by the World Health Organization on March 11, 2020.^1,2^ As of August 1, 2020, over 17 million cases and over 675,000 deaths due to COVID-19 have been reported around the world,^3^ caused by the novel coronavirus severe acute respiratory syndrome coronavirus 2 (SARS-CoV-2).^4,5^ NVX-CoV2373 is a recombinant nanoparticle vaccine constructed from the full-length, wild-type SARS-CoV-2 spike glycoprotein, which mediates viral attachment to the human angiotensin-converting enzyme 2 (hACE2) receptor of host cells for cellular entry and serves as a key target for development of antibodies and vaccines.^6,7^ In rodent and nonhuman primate (NHP) challenge models, NVX-CoV2373/Matrix-M1 induced dose-dependent high titers of antibodies measured against anti-spike protein and hACE2 receptor binding and achieved neutralization of wild-type virus that exceeded the magnitude of responses measured in human convalescent sera and provided protection against SARS-CoV-2 challenge.^8,9^ In addition, CD4+ and CD8+ T-cell responses were induced with a T helper 1 (Th1) dominant phenotype.^8^

We report here on the first part of a two-part, randomized, placebo-controlled, observer-blinded, phase 1/2 trial, which commenced in May 2020 to evaluate the safety and immunogenicity of both 5- and 25-μg doses of NVX-CoV2373 with or without Matrix-M1 adjuvant (50-μg dose) in healthy adults less than 60 years of age. Phase 2 will commence following review of the Day 35 primary safety and immunogenicity analysis of the phase 1 data.

## Methods

### Trial Design and Oversight

Our first-in-human, phase 1 trial evaluated the safety and immunogenicity of NVX-CoV2373 with or without Matrix-M1 adjuvant at two phase 1 clinical units in Australia (Nucleus Network, Melbourne, Victoria). Eligible participants were healthy men and non-pregnant women 18 to 59 years of age with a body mass index 17 to 35 kg/m^2^. Healthy status, assessed during the screening period, was based on medical history and clinical laboratory, vital signs, and physical examination findings. Participants with a history of SARS or COVID-19 or who tested positive at screening (by real-time polymerase chain reaction [RT-PCR] or enzyme-linked immunosorbent assay [ELISA] testing) along with participants exposed to individuals with confirmed SARS-CoV-2 or working in an occupation at high-risk for SARS-CoV-2 exposure were excluded. Details of the trial design, conduct, oversight, and analyses are provided in the protocol and statistical analysis plan (available at NEJM.org). All participants provided written informed consent prior to trial enrollment.

As a safety measure, six participants were initially randomly assigned (1:1) into the two-dose NVX-CoV2373/Matrix-M1 groups, vaccinated in an open-label manner, and observed for reactogenicity through 48 hours. Thereafter, the remaining participants (n=125) were randomly assigned (1:1:1:1:1) in a blinded manner to one of five vaccine groups according to pregenerated randomization schedules without stratification (Fig. 1).

**Figure 1.**
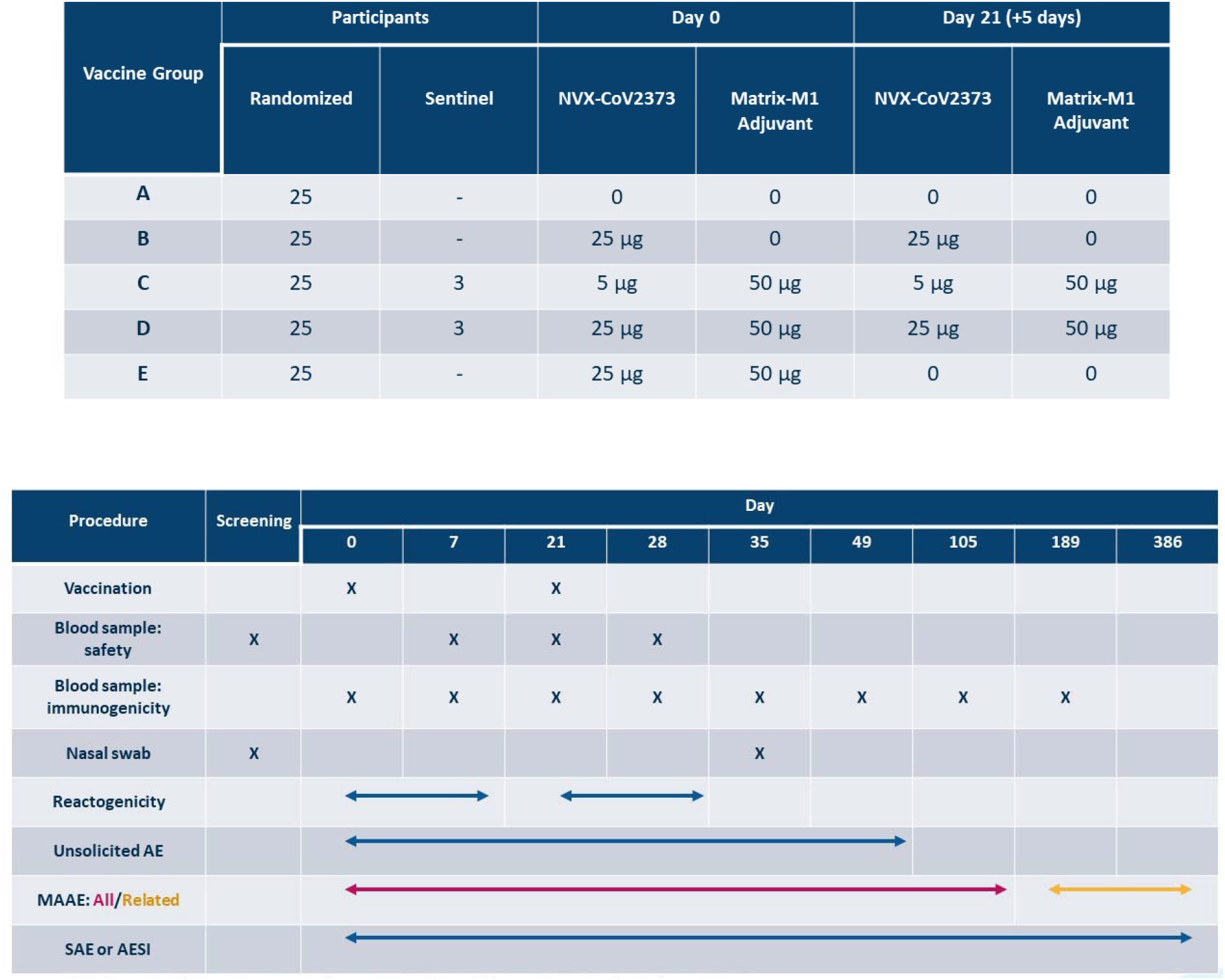
Vaccine Regimens and Key Trial Timings. AESI denotes adverse event of special interest, MAAE medically attended adverse event, SAE serious adverse event.

The trial was designed by Novavax, Inc. (Gaithersburg, MD), with funding support from the Coalition for Epidemic Preparedness Innovations. The trial protocol was approved by the Alfred Hospital Human Research Ethics Committee (Melbourne, Australia) and performed in accordance with the International Conference on Harmonization, Good Clinical Practice guidelines. Safety oversight for specific vaccination pause rules and advancement to phase 2 is supported by an independent safety monitoring committee. The authors assume responsibility for the accuracy and completeness of the data and analyses, as well as for the fidelity of the trial.

### Trial Vaccine, Adjuvant, and Placebo

NVX-CoV2373, developed by Novavax and manufactured at Emergent Biosolutions (Rockville, Maryland), is a recombinant nanoparticle vaccine constructed from the full-length, wild-type SARS-CoV-2 spike glycoprotein (GenBank gene sequence MN908947, nucleotides 21563-25384) optimized in the established baculovirus*-Spodoptera frugiperda* (Sf9) insect cell expression system.^8^ NVX-CoV2373 is generated with 682-QQAQ-685 mutations at the S1/S2 cleavage sites to confer protease resistance and two proline substitutions at residues K986P and V987P at the top of the heptad repeat 1/central helix in the S2 subunit to stabilize the construct in a prefusion conformation. NVX-CoV2373 is resistant to proteolytic cleavage, binds with high affinity to the hACE2 receptor, and demonstrates thermostability.^8,10^ Matrix-M1, a saponin-based adjuvant,^11^ was manufactured at Novavax AB (Uppsala, Sweden). Both vaccine and adjuvant are stored at 2°C to 8°C. Placebo was sterile 0.9% normal saline.

NVX-CoV2373 and Matrix-M1 were mixed at the site, and the injection volumes were 0.6 mL. Each participant received two intramuscular injections of trial vaccine or placebo, one injection on Days 0 and 21, in alternating deltoids. Participants and trial site personnel managing the conduct of the trial remained blinded to vaccine assignment, with the exception of sentinel dosing. Vaccination pause rules were in place to monitor participant safety (Supplementary Appendix).

### Safety Assessments

For the Day 35 primary analysis, safety was assessed across trial visits (Fig. 1) and included short-term reactogenicity, unsolicited adverse events, adverse events of special interest (AESI), clinical laboratory tests, and vital signs.

Participants were observed for at least 30 minutes after each vaccination for assessment of solicited reactogenicity and were asked to continue monitoring these events at home for 7 days after each vaccination using a participant diary. Predefined solicited local (injection site) reactogenicity included pain, tenderness, erythema, and swelling, and systemic reactogenicity included temperature/fever, nausea/vomiting, headache, fatigue, malaise, muscle pain/myalgia, and joint pain/arthralgia. Reactogenicity severity was graded according to standard toxicity criteria (Table S2^12^).

Unsolicited adverse events were assessed for clinical severity (mild, moderate, and severe) and causality (not related and related). Assessments included AESI relevant to COVID-19 (Table S3^13,14^) and potential immune-mediated medical conditions (Table S4). Participants underwent nasopharyngeal swab testing for COVID-19 on Day 35 or any time they reported symptoms suggestive of possible infection. Laboratory safety was graded according to standard toxicity criteria adjusted for local laboratory ranges (Table S5^12,13^); standard toxicity criteria were also applied to vital signs (Table S6^12^).

### Immunogenicity Assessments

For the Day 35 primary analysis reported herein, immunogenicity assessments included an anti-spike immunoglobulin G (IgG) ELISA (Days 0, 7, 21, 28, and 35), a wild-type virus microneutralization assay (MN) with an inhibitory concentration of > 99% (MN IC_>99_) (Days 0, 21, and 35), and intracellular cytokine staining of antigen-specific CD4^+^ T cells (Days 0 and 28). Details of these assays are provided in the Supplementary Appendix. Human ACE2 receptor binding inhibition by ELISA and additional testing of peripheral blood mononuclear cells for T-cell responses are ongoing.

Immunogenicity results were compared to a control panel of 32 convalescent serum specimens from PCR-confirmed COVID-19 from Baylor College of Medicine, which were classified by clinical severity independent from Novavax (Supplementary Appendix and Table S1).

### Statistical Analysis

The sample size for the first-in-human trial was based on clinical and practical considerations, not on a formal statistical power calculation. Most endpoints were summarized using geometric means with 95% confidence intervals based on the t distribution of the log-transformed values.

The primary safety and immunogenicity analysis was conducted after all participants were followed through Day 35. Only sponsor personnel not involved in the conduct of the trial were unblinded at the individual vaccine-group assignment level.

## Results

### Trial Population

The trial initiated on May 26, 2020, with 134 participants randomized from May 27 to June 6, 2020; this includes three participants serving as back-ups for sentinel dosing who immediately discontinued the study prior to being vaccinated (Fig. S1). All randomized participants received their first vaccination on Day 0, and all but three received their second vaccination at least 21 days later. Three participants did not receive the second vaccination; two in the placebo group withdrew consent (unrelated to any adverse event) and one in the 25-μg NVX-CoV2373/Matrix-M1 group had an unsolicited adverse event of mild cellulitis, which started 8 days after vaccination without preceding local symptomatology and was assessed as not related to trial vaccine. Demographic characteristics of the participants are presented in Table 1.

**Table 1.** Demographic Characteristics of the Participants in the NVX-CoV2373 First-in-Human Trial 2019nCoV-101 at Enrollment.

|  | Group | A | B | C | C-Sentinel | D | D-Sentinel | E |  |
| --- | --- | --- | --- | --- | --- | --- | --- | --- | --- |
|  | NVX-CoV2373 Dose 1/2 (µg) | 0/0 | 25/25 | 5/5 | 5/5 | 25/25 | 25/25 | 25/0 |  |
|  | Matrix-M1 Dose 1/2 (µg) | 0/0 | 0/0 | 50/50 | 50/50 | 50/50 | 50/50 | 50/0 | Total |
| Characteristic | N | 23 | 25 | 26 | 3 | 25 | 3 | 26 | 131 |
| Sex – no. (%) |  |  |  |  |  |  |  |  |  |
| Male |  | 11 ( 47.8) | 12 ( 48.0) | 13 ( 50.0) | 2 ( 66.7) | 17 ( 68.0) | 2 ( 66.7) | 9 ( 34.6) | 66 ( 50.4) |
| Female |  | 12 ( 52.2) | 13 ( 52.0) | 13 ( 50.0) | 1 ( 33.3) | 8 ( 32.0) | 1 ( 33.3) | 17 ( 65.4) | 65 ( 49.6) |
| Age – yr* |  | 30.3 (10.92) | 27.2 (9.38) | 29.5 (7.99) | 23.7 (7.37) | 35.6 (12.50) | 25.0 (4.58) | 33.0 (8.91) | 30.8 (10.20) |
| Race or ethnic group – no. (%) |  |  |  |  |  |  |  |  |  |
| American Indian or Alaska Native |  | 1 ( 4.3) | 1 ( 4.0) | 2 ( 7.7) | 0 | 1 ( 4.0) | 0 | 2 ( 7.7) | 7 ( 5.3) |
| Asian |  | 2 ( 8.7) | 0 | 6 ( 23.1) | 1 ( 33.3) | 3 ( 12.0) | 1 ( 33.3) | 4 ( 15.4) | 17 ( 13.0) |
| Black or African American |  | 0 | 0 | 0 | 0 | 1 ( 4.0) | 0 | 1 ( 3.8) | 2 ( 1.5) |
| Multi-racial |  | 1 ( 4.3) | 0 | 0 | 0 | 0 | 0 | 0 | 1 ( 0.8) |
| Native Hawaiian or other Pacific Islander |  | 0 | 0 | 0 | 0 | 0 | 0 | 1 ( 3.8) | 1 ( 0.8) |
| Not Reported |  | 0 | 0 | 0 | 0 | 0 | 0 | 0 | 0 |
| White |  | 19 ( 82.6) | 24 ( 96.0) | 18 ( 69.2) | 2 ( 66.7) | 20 ( 80.0) | 2 ( 66.7) | 18 ( 69.2) | 103 ( 78.6) |
| Hispanic or Latino |  | 2 ( 8.7) | 3 ( 12.0) | 6 ( 23.1) | 0 | 3 ( 12.0) | 0 | 5 ( 19.2) | 19 ( 14.5) |
| Body mass index**† |  | 24.94 (3.418) | 25.59 (4.217) | 24.10 (3.872) | 21.43 (2.401) | 26.18 (3.454) | 25.67 (2.294) | 25.52 (3.342) | 25.19 (3.672) |

\* Plus-minus values are means ± standard deviation.
† The body-mass index is the weight in kilograms divided by the square of the height in meters. This calculation was based on the weight and height measured at the time of screening.

### Safety Outcomes

Overall reactogenicity was generally mild, and vaccinations were well tolerated. Following first vaccination, local reactogenicity was more frequent for the NVX-CoV2373/Matrix-M1 regimens with tenderness and pain being the most common (Fig. 2 and Table S7). Systemic events were individually less frequent, with headache, fatigue, and myalgia being the most common and also observed with greater frequency in the NVX-CoV2373/Matrix-M1 regimens. Following second vaccination, NVX-CoV2373/Matrix-M1 induced greater local and systemic reactogenicity, but the majority of reported symptoms remained at grade ≤1. Only one participant experienced fever (mild) after second vaccination (Day 1 only). Importantly, mean durations of events were ≤2 days without appreciable change in duration with second vaccination. Two participants (one placebo) had reactogenicity that extended beyond Day 7. Severe reactogenicity was infrequent, seen more with systemic symptoms and following second vaccination. In total, two participants had severe events following first vaccination and eight participants following second vaccination, with placebo recipients citing similar frequencies as those receiving NVX-CoV2373 with second vaccination. No participants sought medical intervention or refused second vaccination because of reactogenicity.

**Figure 2.**
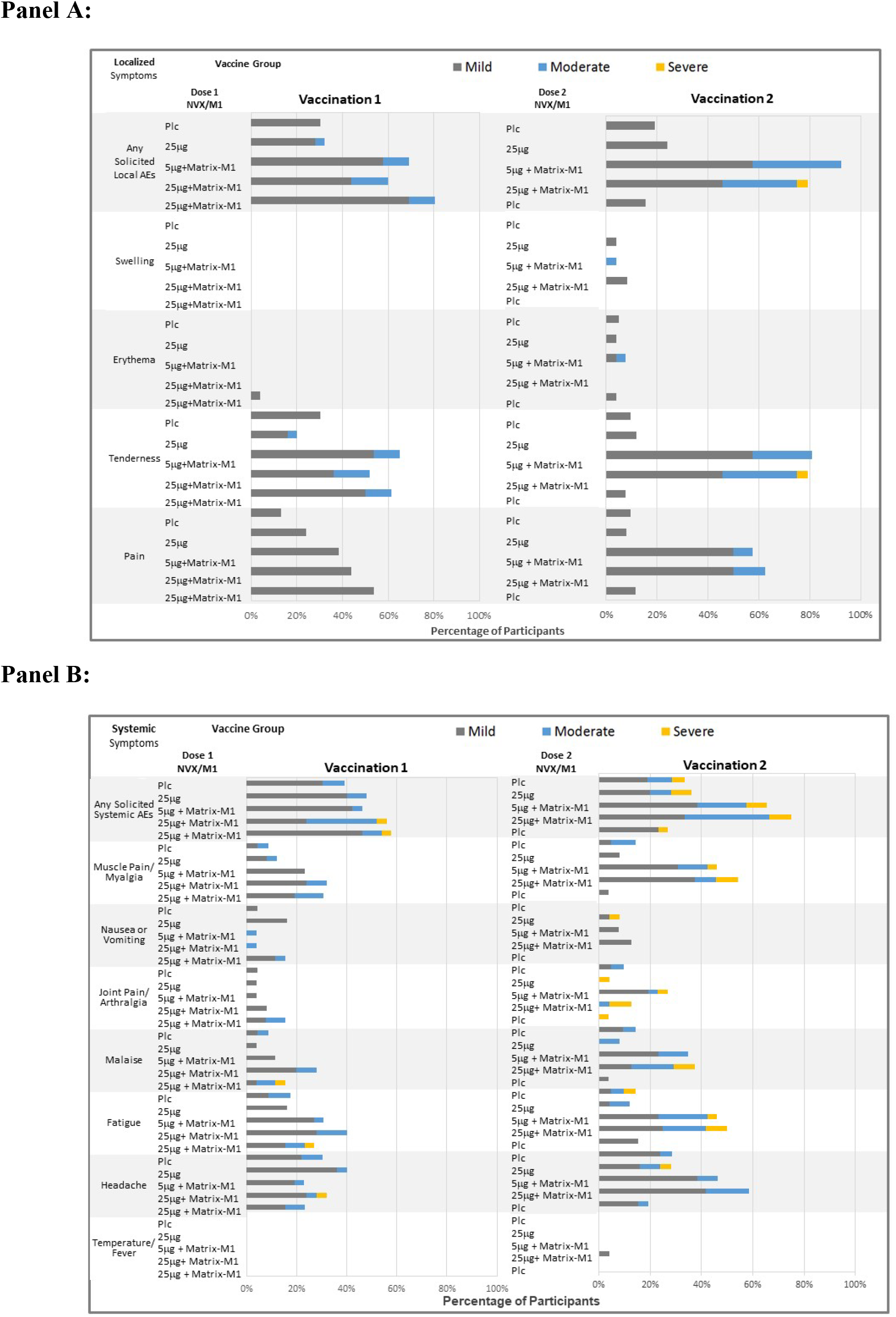
Solicited Local and Systemic Adverse Events. Percentage of participants per vaccine group by maximum reactogenicity grade through 7 days following each vaccination is plotted. There were no Grade 4 (life-threatening) events. Those participants who reported zero events make up the remainder of the 100% calculation (not displayed).

Laboratory abnormalities of grade 2 or higher occurred in very few participants and were not associated with any clinical manifestations or worsening with repeat vaccination (Supplementary Appendix and Table S8). Vital signs remained stable immediately following vaccination and at all visits.

Unsolicited adverse events (Table S9) were mild in nature and occurred at similar frequencies across the active vaccine groups. A slight decrease was observed after second vaccination but attributed to a shorter duration of observation. Outside of administrative site conditions and safety laboratory monitoring, the most frequent was in the system organ classification of infections and infestations. No serious adverse events or AESIs were reported through Day 35, and vaccination pause rules were not implemented.

### Immunogenicity Outcomes

ELISA anti-spike IgG geometric mean ELISA units (GMEUs) were low at Day 0. By Day 21, robust responses occurred for all NVX-CoV2373/Matrix-M1 regimens, with geometric mean fold rises (GMFR) exceeding those induced by unadjuvanted NVX-CoV2373 by at least 10-fold (Fig. 3A and Table S10). Within 7 days following second vaccination with NVX-CoV2373/Matrix-M1 (Day 28), GMEUs increased an additional eight-fold over responses seen with first vaccination and within 14 days (Day 35) responses had more than doubled yet again, achieving GMFRs approximately 100-fold over those observed with NVX-CoV2373 alone. A single vaccination with NVX-CoV2373/Matrix-M1 achieved similar GMEU levels to those in asymptomatic (exposed) COVID-19 patients, while a second vaccination achieved levels that exceeded convalescent serum from outpatient-treated COVID-19 patients by six-fold, achieved levels similar to convalescent serum from patients hospitalized with COVID-19, and exceeded overall convalescent serum GMEUs by nearly six-fold. The responses in the two-dose 5-μg and 25-μg NVX-CoV2373/Matrix-M1 regimens were similar, further highlighting the role of the adjuvant for dose sparing.

**Figure 3.**
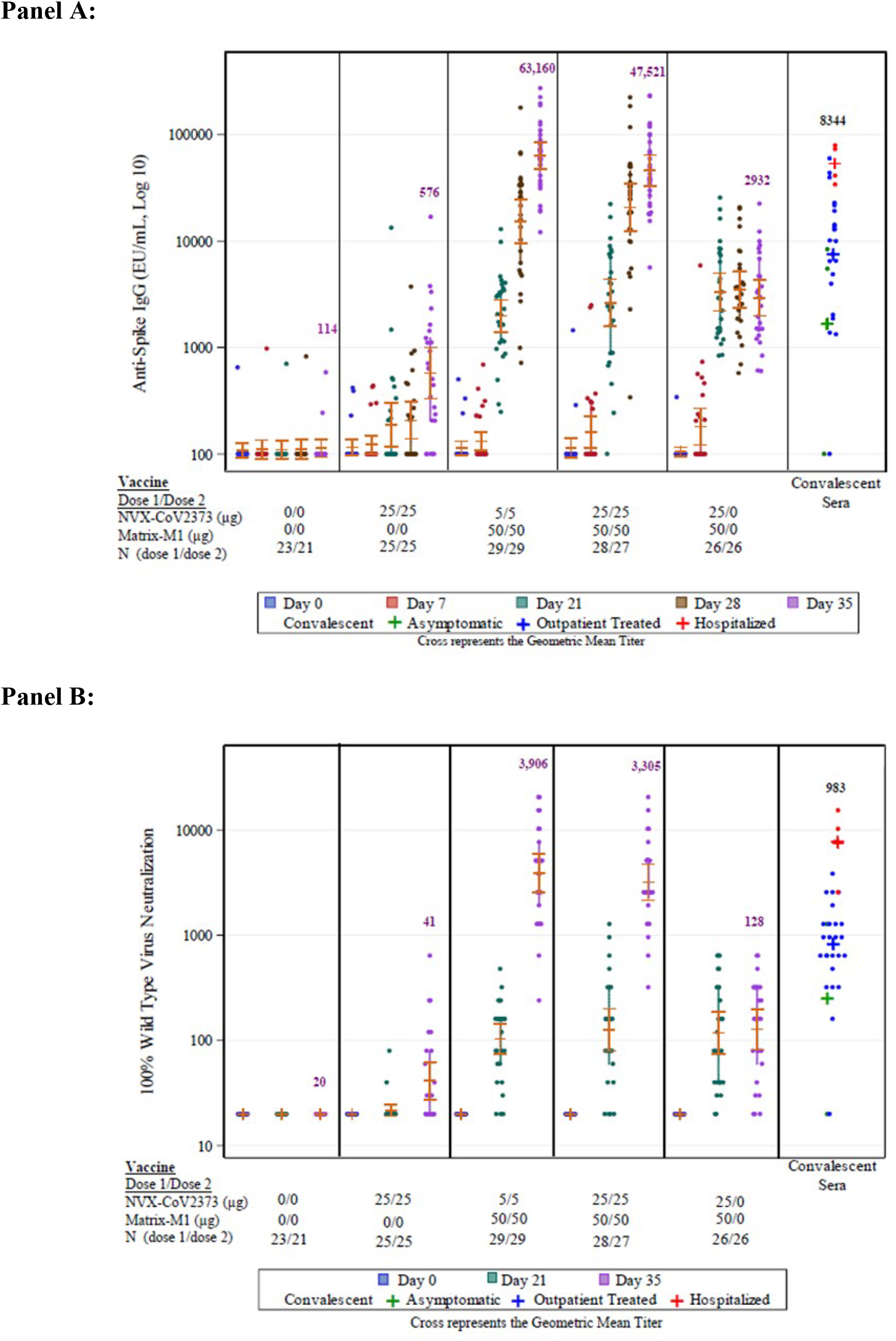
SARS-CoV-2 Anti-Spike IgG and Neutralizing Antibody Responses. Shown are geometric mean anti-spike IgG enzyme-linked immunosorbent assay (ELISA) unit responses to NVX-CoV2373 protein antigens (Panel A) and wild-type SARS-CoV-2 microneutralization assay at an inhibitory concentration > 99% (MN IC_>99_) titer responses (Panel B). Horizontal bars and whisker endpoints represent geometric mean titer (GMT) values and their 95% confidence intervals, respectively. The convalescent serum panel includes specimens from PCR-confirmed COVID-19 participants from Baylor College of Medicine (29 specimens for ELISA and 32 specimens for MN IC_>99_). Severity of COVID-19 is denoted as a red mark for hospitalized patients (including intensive care setting), a blue mark for outpatient-treated patients (sample collected in emergency department), and a green mark for asymptomatic (exposed) patients (sample collected from contact/exposure assessment).

Neutralizing antibodies were undetectable prior to vaccination and had similar patterns of response following vaccination with MN IC_>99_ titers most pronounced for the NVX-CoV2373/Matrix-M1 regimens (Fig. 3B and Table S11). Following first vaccination, GMFR were approximately five-fold greater with NVX-CoV2373/Matrix-M1 than NVX-CoV2373 alone. Second vaccinations with adjuvant had a profound effect on MN titers – inducing >100-fold rise over single vaccinations without adjuvant. When compared to convalescent serum, second vaccinations with NVX-CoV2373/Matrix-M1 achieved GMT levels four-fold greater than outpatient-treated COVID-19 patients, levels spanning those of patients hospitalized with COVID-19, and exceeded overall convalescent serum GMT by four-fold.

Convalescent serum, obtained from COVID-19 patients with clinical symptoms requiring medical care, demonstrated proportional IgG and MN immune responses that increased with illness severity (Fig. 3).

A strong correlation was observed between neutralizing antibody titers and anti-spike IgG (r=0.9466, Fig. 4), which was not observed with unadjuvanted vaccine (r=0.7616), but was similar to that of convalescent sera (r=0.958). Both 5- and 25-μg NVX-CoV2373/Matrix-M1 demonstrated similar magnitudes of two-dose responses and every participant seroconverted using either assay measurement when a two-dose regimen was utilized.

**Figure 4.**
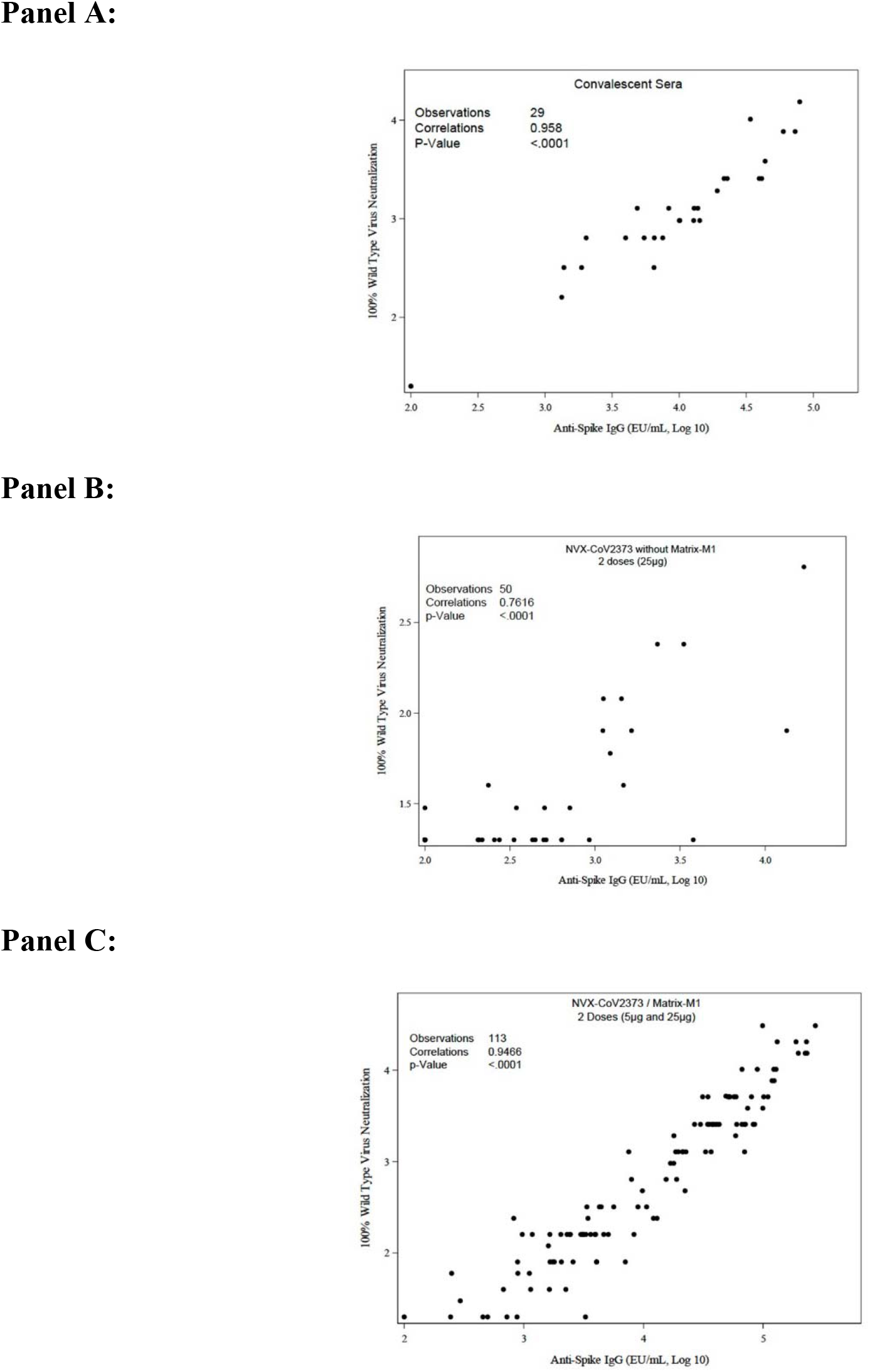
Correlations of Anti-Spike IgG and Neutralizing Antibody Responses. Shown are scatter plots of 100% wild-type neutralizing antibody responses and anti-spike IgG ELISA unit responses for convalescent comparator serum (Panel A), two-dose 25-ng unadjuvanted vaccine (Panel B), and two-dose 5- and 25-μg adjuvanted vaccine (Panel C).

T-cell responses in 16 participants (randomly selected from Groups A through D) showed that NVX-CoV2373/Matrix-M1 regimens induced antigen-specific polyfunctional CD4+ T-cell responses in terms of IFN-γ IL-2, and TNF-α production upon spike protein stimulation. There was a strong bias toward this Th1 phenotype (Fig. 5).

**Figure 5.**
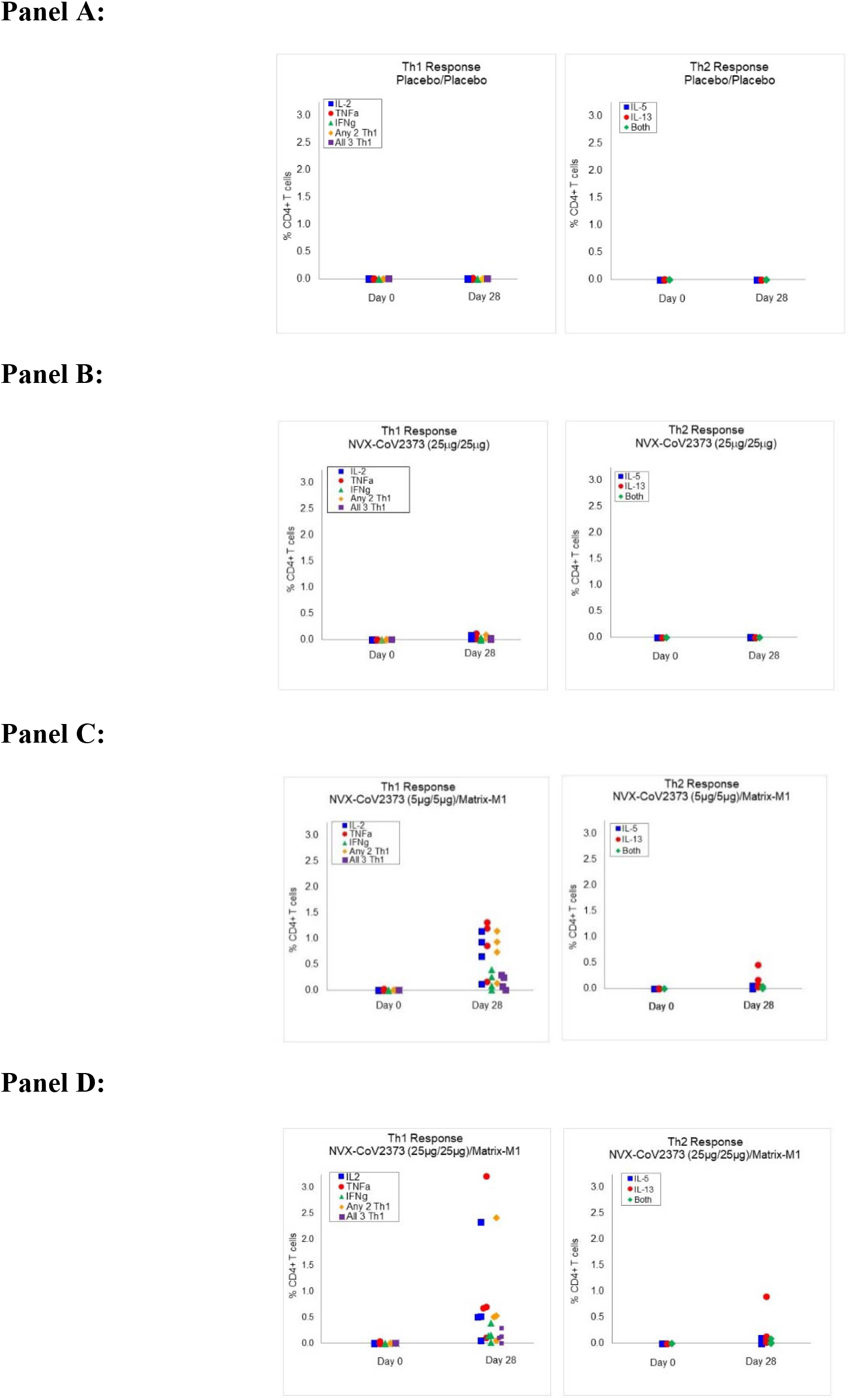
NVX-CoV2373 CD4^+^T-cell Responses. Frequencies of antigen-specific CD4^+^T cells producing T helper 1 (Thl) cytokines interferon-gamma (IFN-γ), tumor necrosis factor-alpha (TNF-α), and interleukin (IL)-2 and for T helper 2 (Th2) cytokines IL-5 and IL-13 indicated cytokines from 4 participants each in Groups A, B, C, and D following stimulation with the recombinant spike protein. “Any 2” in Thl cytokine panel means CD4^+^T cells that can produce two types of Thl cytokines at the same time. “All 3” indicates CD4^+^ T cells that produce IFN-γ, TNF-α, and IL-2 simultaneously. “Both” in Th2 panel means CD4^+^ T cells that can produce Th2 cytokines IL-5 and IL-13 at the same time.

## Discussion

The primary safety and immunogenicity analyses indicate that in healthy adult participants 18 to 59 years of age, the two-dose regimens of 5- and 25-μg NVX-CoV2373/Matrix-M1 were well tolerated and induced the most robust immune responses with high levels of neutralizing antibodies that closely correlated with anti-spike IgG. Furthermore, neutralizing antibody responses following second vaccination were of the magnitude seen in convalescent serum from hospitalized COVID-19 patients and exceeded overall convalescent sera GMT by four-fold. The benefit of Matrix-M1 adjuvant was clear in the magnitude of the antibody and T-cell response, induction of functional antibodies, and dose sparing.

Although the effector mechanisms engaged for protection with a COVID-19 vaccine are yet not known, it is widely held that neutralizing antibodies are likely to be associated with protection and that their concentration may drive the level of efficacy, but the levels of neutralizing antibodies that will confer protection are not known at this time. In the FDA guidance for use of convalescent serum antibody for therapy, the Agency made the reasonable presumption that sera with a MN titer of at least 1:160 should be used and this guidance reflects the consensus that SARS-CoV-2 specific MN are likely to be associated with protection.^15^ Convalescent plasma treatments have shown promising results in seriously ill COVID-19 patients using plasma with neutralization titers in the range of 1:80 to 1:640, with some dose-dependent observations.^16-20^ The convalescent serum comparator used in our assays largely reflects patients who experienced clinical symptoms requiring medical care. Interestingly, in convalescent serum, spike IgG and MN titers stratified by illness severity reflect findings in other studies with COVID-19 where illness severity and the magnitude of MN responses are proportional. Although these findings have to be cautiously interpreted, they align with the general findings in viral respiratory disease, particularly those infecting via class I fusion proteins such as RSV, that MN responses induced by infections are associated with protection.^21^

Notably in NHP studies, clinical doses of vaccine (5- and 25-μg NVX-CoV2373/Matrix-M1) resulted in sterile immunity in the lungs and nasal passage following wild-type virus challenge, suggesting that the vaccine may both protect against upper and lower respiratory tract disease and interrupt transmission (Smith, submitted).

Novavax has accumulated safety on over 14,000 subjects in various nanoparticle vaccine trials, including children, pregnant women and older adults^22-25^ and over 4,200 subjects exposed to Matrix-M1 adjuvant (5 months to 85 years of age). The current study demonstrates a safety profile that is acceptable and comparable to past studies with these platform technologies. Overall, mild reactogenicity was noted by the majority of subjects with an adjuvanted vaccine slightly increasing the frequency and intensity of these events. These safety findings are consistent with our seasonal influenza program currently in late stage clinical development.^25^

The value of the second dose of the two-dose regimen is very high levels of antibodies that warrant the logistics of a boosting visit. Additionally, the vaccine is a liquid formulation that can be stored at 2°C to 8°C, allowing for successful cold chain management with existing infrastructure.

Trial limitations include the length of time of safety observations and measured immune responses, as safety in the face of exposure to virus infection and durability of responses for SARs-CoV-2 vaccines will be of great interest. Previous experience with a full-length Ebola glycoprotein nanoparticle vaccine adjuvanted with Matrix-M1 suggests that vaccine-specific immune responses may be detected out to 1 year.^26^ Although in recent nonclinical studies (Smith, submitted),^8^ animals immunized and challenged with live virus did not exhibit features of vaccine-enhanced disease, careful safety monitoring in the context of human efficacy trials will need to be implemented. Additionally, this trial was performed in a healthy population of adults aged 18-59 years of age without significant comorbid disease or known immunocompromising conditions. The populations at greatest risk of serious COVID-19 include people with comorbidities and older adults, and planned vaccine evaluations will address these populations. Notably, recent clinical trials evaluating a recombinant seasonal influenza hemagglutinin nanoparticle vaccine adjuvanted with Matrix-M1 in nearly 2,300 adults 65 years of age or older with comorbidities suggested that the combination of Matrix-M1 with nanoparticle vaccine were both safe and could induce robust functional antibody and T-cell responses in older adults including those with a modest level of comorbidities.^25,27^

Taken together, the adjuvanted, recombinant, full-length spike protein nanoparticle vaccine is a promising candidate that warrants rapid advancement into efficacy studies.

## Data Availability

These are interim data, and individual participants remain masked to individual vaccine assignment. Therefore, it would be inappropriate to share individual level results at this time.

